# An observational study of the association between COVID-19 vaccination rates and entry into the Australian ‘Million Dollar Vax’ competition

**DOI:** 10.1101/2022.03.02.22271734

**Authors:** Dajung Jun, Anthony Scott

## Abstract

**Objectives:** To examine the association between financial incentives from entry into a vaccine competition with the probability of vaccination for COVID-19.

**Design:** A cross-sectional study with adjustment for covariates using logistic regression

**Setting:** October and November 2021, Australia.

**Participants:** 2,375 respondents of the Taking the Pulse of the Nation Survey

**Primary and secondary outcome measures:** The proportion of respondents who had any vaccination, a first dose only, or second dose after the competition opened.

**Results:** Those who entered the competition were 2.27 (95% CI 1.73 to 2.99) times more likely to be vaccinated after the competition opened on October 1^st^ than those who did not enter—an increase in the probability of having any dose of 0.16 (95 % CI 0.10 to 0.21) percentage points. This increase was mostly driven by those receiving second doses. Entrants were 2.39 (95% CI 1.80 to 3.17) times more likely to receive their second dose after the competition opened.

**Conclusions:** Those who entered the Million Dollar Vax competition were more likely to receive a vaccination after the competition opened compared to those who did not enter the competition, with this effect dominated by those receiving second doses.

**Strengths and limitations of this study:**

- We use a nationally representative sample of individual self-reported vaccination status and timings.
- We distinguish between the association between competition entry and first and second doses.
- We adjust for a rich set of individual characteristics associated with vaccination status, and examine the factors influencing competition entry
- The strong association for second dose vaccinations may reflect some individuals who had already scheduled their second dose after the competition opened, potentially leading to an overestimate of the association.

## Background

The effectiveness of using financial incentives to increase vaccination rates for the SARS-COV-2 virus is uncertain.^[1],[2],[3]^ One form of financial incentive has been the entry into vaccination competitions where participants are eligible for large randomly-drawn cash prizes. These have also been referred to as lotteries but unlike lotteries, they do not require cash payment on entry and are not a form of profit-driven gambling. Financial incentives have been used before to encourage childhood immunisation, but not in the form of competitions with cash prizes. Such competitions were established in 2021 to increase vaccination rates for COVID-19, mostly in the United States. For example, the competition in Ohio was run from May to June 2021 with 5 x $1 million prizes over five weeks.

However, the evidence on the effect of such competitions on vaccination rates has been mixed. Four studies using state-level data on vaccination rates over time, and comparing states with vaccination competitions with those with none, found they were ineffective in increasing vaccination rates.^[4],[5],[6],[7]^ Four studies found an increase in vaccination rates^[8], [9], [10], [11]^, including one that found increases in vaccination rates in low-income counties in Ohio but not in high-income counties.^[9]^ One study examined the use of financial incentives across 24 states across the U.S., mainly including vaccination competitions, and found no overall impact on vaccination rates.^[12]^ The reason for these mixed results is unclear as all used aggregate state-level data on changes in vaccination rates over time through each used slightly different methodologies.

Unlike most of this previous work, the aim of this research is to conduct a more granular analysis using individual-level data to examine the association between an individual’s decision to get vaccinated and financial incentives. The Million Dollar Vaccination Campaign (M$V) was open to entries from 1^st^ to 31^st^ of October 2021 for those aged 18 years or over who were Australian residents. This was accompanied by a significant national marketing campaign that specifically targeted local areas with low vaccination rates and with populations finding it difficult to access vaccinations. If an entrant was chosen to receive a prize, they were required to show proof of two-dose vaccination in the form of a government-approved electronic vaccination certificate.

M$V was funded by an alliance of philanthropic organisations coordinated by the Summer Foundation. The competition was designed to increase the rate of full (two-dose) vaccinations in the context of meeting national vaccination targets that would trigger the end of harsh lockdowns in the two most populous states, New South Wales and Victoria. The objective was to speed up the rate of vaccination amongst those who intended to get vaccinated but had not yet done so. This was intended to reduce hospitalisations and ongoing economic costs of lockdowns. Australia’s vaccination program started in March 2021. The Therapeutic Goods Administrative approved three vaccines for Australians’ use in 2021: Pfizer, AstraZeneca, and Moderna, each requiring two doses for ‘full vaccination’. On the 30^th^ of September, just before the competition opened, vaccination rates had steadily increased to 77.8% percent of the population over 16 years old with a first dose and 54.2% with a second dose. New South Wales (NSW) and Victoria, the two most populous states, had experienced outbreaks since July 2021 and were under various forms of lockdown at the end of September, including night-time curfews in Victoria, closure of retail businesses and hospitality, and continuing bans on travel. Lockdowns in NSW were more targeted at specific Local Government Areas (LGAs) with high case numbers. All eight states and territories agreed to a national roadmap on 6^th^ August 2021, with states individually releasing precise targets of population vaccination rates that were linked to the lifting of restrictions throughout the last quarter of 2021, with some target dates at the time the competition was open. For example, in Victoria, the targets were 70% of the population aged 16 and over, (reached on 21^st^ October), 80% (reached on 29^th^ October), and 90% of 12+ years (reached on 18^th^ November) with a second dose. These targets provided non-financial incentives to get a second dose (referred to as fully vaccinated at the time) as restrictions were eased when targets were met, with restrictions largely non-existent after the 90% target was reached.

The competition provided the potential to receive financial incentives to encourage receipt of the first dose for those not vaccinated and provided incentives to those with a first dose to schedule a second dose if they had not already done so. The interval between the first and second doses at the time depended on the vaccine: 4-8 weeks for Astra Zeneca during an outbreak (up to 12 weeks with no outbreak) and 3-6 weeks for Pfizer from July 2021.^[13]^ Those with a first dose may already have had their second dose scheduled during October given the recommended fixed interval between doses, and so the incentives would not influence this group unless they changed their scheduled appointment to receive their second dose earlier or were persuaded not to delay their appointment. Those who already had their second dose before the competition opened could still enter, but their vaccination status would not be affected by the competition.

## Method

### Patient and public involvement statement

There was no patient or public involvement in the research.

### Data and participants

The Taking the Pulse of the Nation (TTPN) Survey was run by the Melbourne Institute and was administered every week from April 2020 and every two weeks from January 2021. Each wave included 1,200 different respondents and so is a repeated cross-section design. The analysis used data from 2,400 respondents in Waves 44 and 45 conducted in November 2021 after the competition was closed at the end of October. Of 2,400 respondents, 2,375 responded to the vaccination question. A further 13 respondents did not know the month they received their first vaccination, leaving 2,362 for our analysis as a final sample.

The TTPN Survey dataset was collected by a commercial provider using a mixed-mode procedure. TTPN was designed to track changes in the economic and social well-being of Australians during the pandemic. For each wave, 400 respondents were interviewed by telephone, and 800 respondents completed a web survey. The survey provider constructed the sampling frame from a diverse set of continuously updated proprietary databases. The survey sampling procedure followed strict quotas for six states and the Australian Capital Territory (ACT). Each wave included 600 men and 600 women, and the shares of respondents for each state and ACT are proportional to the population of that state or territory. Data collection for each survey wave took up to six days to collect until the gender/state quotas are reached.

These data have been extensively used in previous research about COVID-19 including Australian’s hesitancy to get vaccinated, vaccine choice, border re-opening decisions, and responses towards workplace vaccination and testing mandate.^[14], [15], [16], [17]^

The raw share of each state/location/gender/age-group strata in the survey sample was not necessarily the same as the share of this stratum in the population. For each survey wave, post-stratification inverse probability weights were calculated based on Greater Capital City Statistical Area (GCCSA) or ‘Rest of State’ for each state using respondents’ postcode, age group (18-24, 24-35, 35-44, 45-54, 55-64, 64-75), and gender. The populations of each stratum are calculated based on the latest ABS estimated resident population projections from the 2016 Census. These weights were used in all analyses.

### Study design and hypothesis

Using data from a cross-sectional survey, the main hypothesis is whether the proportion of all respondents who were vaccinated after September 30^th^ is different for those who entered the competition compared to those who did not. Unlike some U.S. lotteries where the whole population was automatically entered, each person entered the M$V voluntarily by completing a short webform providing their contact details. Proof of vaccination was not required at entry though individuals had to tick a box on the webpage stating that they had at least their first dose. Those who had already had their second dose before the competition opened could enter. If they were chosen to receive a prize (a provisional winner), they were required to show proof of full vaccination (interpreted at the time as two doses) in the form of a government-approved electronic vaccination certificate. To claim a prize full (two-dose) vaccination must have occurred before 13^th^ December, or no later than 13^th^ January, depending on the required interval between first and second doses, which may vary across States and be up to 12 weeks. Only one entry per person was allowed.

The competition had a $AU 1 million ($US 0.72 million) Grand Prize in cash and a total of 3,100 daily prizes of $AU 1,000, with a total prize pool of $AU 4.1 million. Each entrant was eligible for the Grand Prize draw and the daily draw on the entry date. The daily prizes were in the form of a gift card that could be used at a range of participating stores. The lottery was accompanied by a $AU 3 million marketing campaign led by Sayers that included peak-time TV, radio, and full-page national and regional newspaper advertising, extensive social media advertising, and outdoor media. The campaign targeted culturally and linguistically diverse audiences and included advertising in languages such as Mandarin, Arabic, and Vietnamese, and areas with high populations of Indigenous people. As the campaign progressed, the targeting became more granular and nuanced in response to the analysis of data regarding the reach of the campaign, competition entrants, and vaccination rates in specific geographic locations throughout Australia. In response to concerns raised on social media about M$V being a scam, the campaign pivoted to engage and profile daily draw winners and to provide social proof about the legitimacy of M$V. When the competition closed, 2,744,974 Australians had entered, representing 13.7% of the adult population. The study design exploited information on the month individuals received their first or second dose of a COVID vaccine which was asked in Waves 44 and 45 after the competition had closed.

### Variables

Participants were asked the following questions during Waves 44 and 45 in November 2021 to determine their vaccination status. *“Are you willing to have the COVID-19 vaccine? (1) Yes, (2) No, (3) Don’t Know (4), I have had the first dose of the vaccine only (5), I have had the first and second dose of the vaccine*.” If they answered option (4) they were asked the month of their first vaccination. If they answered option (5), they were asked the month of their first and second vaccination. They were separately asked, *“Did you enter the Million Dollar Vax Lottery? (1) Yes, 2) No*.” which is used to define the main independent variable of competition entry.

The main outcome variable is defined according to the timing of each individual’s vaccination and is equal to one for those who reported receiving any vaccination after the competition opened in October and is zero for the rest of the respondents. The denominator includes respondents who were either unvaccinated or those who received their first or second dose before October. The unvaccinated are in the denominator only for the group who did not enter the competition as this group could potentially have changed their decision in response to the competition – that is they were ‘eligible’ to be vaccinated. In addition, we separately analysed those who had only their first dose after the competition opened and those who had their second dose after the competition opened.

TTPN asked a range of questions known to be associated with vaccination status, so these were included as independent variables in the analysis. We included indicators for male, age categories (aged 25-34; aged 35-44; aged 45-49; aged 50-54; aged55-64; aged 65-74; 75+), having a child under 18, income categories (25-50 percentile; 50-75 percentile; 75 percentile+; refused to report), education categories (high school graduates; some college; university and above), and categories of the industry relative to the unemployed (agriculture; mining; manufacturing; electricity; construction; wholesale; retail; food services; transport; information media; insurance services; real estate services; professional, scientific and technical services; administrative services; public administration; education; healthcare assistance; arts and recreation services; other). These categories are defined using 2006 Australian and New Zealand Standard Industry Classification from the Australian Bureau of Statistics. Indicators for the states of residence and living in a rural area were included. Indicators for financial stress, policy satisfaction (satisfied; not satisfied), voting preferences (liberal or national; labour; greens or democrats) were included, and an indicator for wave 45 (15 - 19, November) was included.

The vaccination rates of individuals could be associated with the vaccination rates of others in their LGA through neighbourhood peer effects, the location of vaccination providers, and other LGA-specific factors. In addition, M$V targeted LGAs with low vaccination rates, and so LGA vaccination rates would be associated with the competition entry. We, therefore, merged data on LGA-level vaccination rates using each respondent’s postcode of residence.

### Statistical analysis

Data were analysed using logistic regression with aforementioned covariates as independent variables to adjust for observed differences between those participating in the competition and those who did not. We chose a logistic model to estimate the probability of receiving a vaccine if the respondent entered the competition after September 30^th^. Separate regressions were conducted for those receiving their first vaccination after September 30^th^ and those receiving their second vaccination after September 30^th^. Results were reported as odds ratios and differences in predicted probabilities of being vaccinated, with 95% confidence intervals.

## Results

### Descriptive statistics of our final sample

When the survey was completed in November (after entry had closed), 60.4% of all respondents had received two doses, and 6.1% had only their first. Among those who had not yet received their first dose, 65.7% were willing to be vaccinated, 21.8% were unwilling to be vaccinated, and 12.4% were unsure.

Table 1 shows the weighted descriptive statistics of the sample used in the analysis and compares those who participated in the competition with those who did not. Seventeen percent of respondents participated in the competition. After the competition opened on October 1^st^, 25.2% of respondents received a vaccination. Of those who entered the lottery, 39.3% received a vaccination after the competition opened on October 1^st^, compared to 22.4% of those who did not enter. After the competition opened, 8.8% of respondents received their first dose. The percentage of those who entered the competition and who received their first dose after it opened was 11.5%, compared to 8.2% for those who did not enter. The proportion who received their second dose after the competition opened was higher at 20.9%. Of those who entered the competition, 34.3% received their second dose after the competition opened compared to 18.2% of respondents who did not enter. Appendix Table A1 shows the unweighted number of respondents in each of the categories of vaccination timing and competition entry which were used to construct the dependent variables in the last three rows of this table.

**Table 1:** Descriptive Statistics.

|  | Full Sample |  | Entrant |  | Non-entrant |  |
| --- | --- | --- | --- | --- | --- | --- |
|  | Mean | Std. | Mean | Std. | Mean | Std. |
| Proportion receiving any dose after September 30th | 0.252 | 0.434 | 0.393 | 0.489 | 0.224 | 0.417 |
| Proportion receiving first dose after September 30th | 0.088 | 0.283 | 0.115 | 0.320 | 0.082 | 0.275 |
| Proportion receiving second dose after September 30th | 0.209 | 0.407 | 0.343 | 0.475 | 0.182 | 0.386 |
| Competition entrant | 0.169 | 0.375 | 1.000 | 0.000 | 0.000 | 0.000 |
| Male | 0.485 | 0.500 | 0.412 | 0.493 | 0.500 | 0.500 |
| Age 18 - 24 | 0.116 | 0.321 | 0.099 | 0.299 | 0.120 | 0.325 |
| Age 25 - 34 | 0.192 | 0.394 | 0.182 | 0.386 | 0.194 | 0.396 |
| Age 35 - 44 | 0.173 | 0.378 | 0.189 | 0.392 | 0.169 | 0.375 |
| Age 45 - 49 | 0.084 | 0.277 | 0.104 | 0.305 | 0.080 | 0.271 |
| Age 50 - 54 | 0.081 | 0.273 | 0.129 | 0.335 | 0.071 | 0.257 |
| Age 55 - 64 | 0.153 | 0.360 | 0.194 | 0.396 | 0.144 | 0.351 |
| Age 65 - 74 | 0.120 | 0.325 | 0.086 | 0.281 | 0.126 | 0.332 |
| Age 75 and above | 0.082 | 0.274 | 0.017 | 0.131 | 0.095 | 0.293 |
| Having a child below 18 | 0.311 | 0.463 | 0.321 | 0.467 | 0.309 | 0.462 |
| Not graduated high school/NA | 0.161 | 0.368 | 0.140 | 0.347 | 0.166 | 0.372 |
| High school graduated | 0.173 | 0.378 | 0.147 | 0.355 | 0.178 | 0.383 |
| Some college | 0.308 | 0.462 | 0.327 | 0.470 | 0.304 | 0.460 |
| University and above | 0.357 | 0.479 | 0.386 | 0.487 | 0.352 | 0.478 |
| Income: below 25 percentile | 0.188 | 0.391 | 0.132 | 0.339 | 0.199 | 0.399 |
| Income: 25 - 50 percentile | 0.288 | 0.453 | 0.281 | 0.450 | 0.290 | 0.454 |
| Income: 50 - 75 percentile | 0.251 | 0.434 | 0.250 | 0.434 | 0.251 | 0.434 |
| Income: 75 and above percentile | 0.199 | 0.400 | 0.235 | 0.425 | 0.192 | 0.394 |
| Income: refused | 0.074 | 0.262 | 0.101 | 0.302 | 0.068 | 0.253 |
| Industry: agriculture, forestry and fishing | 0.014 | 0.119 | 0.011 | 0.105 | 0.015 | 0.122 |
| Industry: mining | 0.008 | 0.089 | 0.011 | 0.105 | 0.007 | 0.085 |
| Industry: manufacturing | 0.026 | 0.159 | 0.021 | 0.144 | 0.027 | 0.162 |
| Industry: electricity, gas, water service | 0.013 | 0.114 | 0.003 | 0.052 | 0.015 | 0.123 |
| Industry: construction and wholesale | 0.043 | 0.202 | 0.051 | 0.220 | 0.041 | 0.198 |
| Industry: retail trade | 0.072 | 0.258 | 0.093 | 0.291 | 0.067 | 0.250 |
| Industry: accommodation and food | 0.021 | 0.143 | 0.014 | 0.119 | 0.022 | 0.148 |
| Industry: transport and warehousing | 0.029 | 0.167 | 0.009 | 0.096 | 0.032 | 0.177 |
| Industry: media and telecommunication | 0.026 | 0.158 | 0.026 | 0.159 | 0.026 | 0.158 |
| Industry: financial and insurance services | 0.044 | 0.205 | 0.028 | 0.164 | 0.047 | 0.212 |
| Industry: rental, hiring and real estate | 0.009 | 0.093 | 0.007 | 0.080 | 0.009 | 0.095 |
| Industry: professional and scientific | 0.043 | 0.203 | 0.045 | 0.208 | 0.043 | 0.202 |
| Industry: administrative and support | 0.019 | 0.138 | 0.021 | 0.142 | 0.019 | 0.137 |
| Industry: public administration and safety | 0.022 | 0.146 | 0.033 | 0.178 | 0.020 | 0.138 |
| Industry: education and training | 0.039 | 0.194 | 0.053 | 0.224 | 0.036 | 0.187 |
| Industry: health care and social assistance | 0.061 | 0.239 | 0.079 | 0.271 | 0.057 | 0.232 |
| Industry: arts and recreation services | 0.011 | 0.105 | 0.016 | 0.127 | 0.010 | 0.100 |
| Industry: other services | 0.059 | 0.235 | 0.054 | 0.226 | 0.060 | 0.237 |
| Industry: refused/don't know/not in the labor force | 0.442 | 0.497 | 0.424 | 0.495 | 0.446 | 0.497 |
| Living in rural | 0.316 | 0.465 | 0.306 | 0.461 | 0.318 | 0.466 |
| NSW | 0.329 | 0.470 | 0.271 | 0.445 | 0.341 | 0.474 |
| VIC | 0.263 | 0.441 | 0.324 | 0.468 | 0.251 | 0.434 |
| QLD | 0.204 | 0.403 | 0.196 | 0.397 | 0.205 | 0.404 |
| SA | 0.070 | 0.255 | 0.057 | 0.232 | 0.073 | 0.260 |
| WA | 0.102 | 0.303 | 0.127 | 0.334 | 0.097 | 0.296 |
| ACT, TAS, NT | 0.031 | 0.174 | 0.025 | 0.157 | 0.033 | 0.178 |
| Fully Vaccinated rate by LGA | 78.420 | 14.000 | 79.794 | 11.624 | 78.166 | 14.424 |
| With Financial Stress | 0.436 | 0.496 | 0.447 | 0.498 | 0.434 | 0.496 |
| Satisfied with policy | 0.428 | 0.495 | 0.435 | 0.496 | 0.427 | 0.495 |
| Not satisfied with policy | 0.252 | 0.434 | 0.211 | 0.409 | 0.260 | 0.439 |
| Indifferent with policy | 0.320 | 0.467 | 0.354 | 0.479 | 0.313 | 0.464 |
| Voting liberal or national | 0.342 | 0.475 | 0.329 | 0.470 | 0.345 | 0.476 |
| Voting labour | 0.324 | 0.468 | 0.350 | 0.478 | 0.319 | 0.466 |
| Voting greens or democrats | 0.114 | 0.318 | 0.086 | 0.281 | 0.120 | 0.325 |
| Voting others/no preference | 0.219 | 0.414 | 0.235 | 0.424 | 0.216 | 0.412 |
| Wave 44 (1 - 6, Nov 2021) | 0.500 | 0.500 | 0.465 | 0.499 | 0.507 | 0.500 |
| Wave 45 (15 - 19, Nov 2021) | 0.500 | 0.500 | 0.535 | 0.499 | 0.493 | 0.500 |
| Number of observations | 2,362 |  | 436 |  | 1,926 |  |

Note: Data are weighted

Those who chose to enter the competition were more likely to be female, more likely to be between 50 and 64 years old, and less likely to be over 65. Those who entered were likely to have a higher income. There was also a higher proportion of entrants in Victoria.

### Regression results for the association of competition entry and vaccination take-up

Table 2 presents the results from the unadjusted logistic regressions that include only the dummy variable (entrants vs non-entrants) as an independent variable, and from the adjusted logistic regressions that include all covariates in Table 1 as independent variables. The differences between the adjusted and unadjusted models are small. Competition entry is associated with a higher proportion of respondents having any dose after September 30^th^. Those who entered were 2.27 times more likely to have a vaccination after September 30^th^ compared to everyone else. This is equivalent to an increase in the probability of having any dose of 0.155 (95% CI 0.100 to 0.210) compared to everyone else. Entry was associated with a 0.022 (95% CI -0.011 to 0.056) increase in the probability of getting the first dose after September 30^th^, but this was not statistically significant in the adjusted analysis, with the association driven by people getting their second dose. Those who entered were 2.39 times more likely to have a second dose after September 30^th^ compared to everyone else. This is equivalent to an increase in the probability of a second dose after September 30^th^ of 0.152 (95% CI 0.098 to 0.206) compared to everyone else.

**Table 2:** Adjusted and Unadjusted Regressions.

|  | Any dose after<br>September 30<sup>th</sup> | First dose after<br>September 30<sup>th</sup> | Second dose after<br>September 30<sup>th</sup> |
| --- | --- | --- | --- |
| **Adjusted analysis** |  |  |  |
| Entrant vs. non-entrant<br>(Odds Ratio, 95% CI) | 2.274***<br>(1.727 to 2.994) | 1.341<br>(0.884 to 2.033) | 2.389***<br>(1.800 to 3.169) |
| Change in probability<br>(95% CI) | 0.155***<br>(0.100 to 0.210) | 0.022<br>(-0.011 to 0.056) | 0.152***<br>(0.098 to 0.206) |
| **Unadjusted analysis** |  |  |  |
| Entrant vs. non-entrant<br>(Odds Ratio, 95% CI) | 2.249***<br>(1.732 to 2.919) | 1.451*<br>(0.971 to 2.169) | 2.351***<br>(1.795 to 3.080) |
| Change in probability<br>(95% CI) | 0.169***<br>(0.111 to 0.228) | 0.033<br>(-0.006 to 0.072) | 0.161<br>(0.105 to 0.217) |
| Number of observations | 2,362 | 2,362 | 2,362 |

Notes: Results are based on logistic regressions and are all weighted. Respondents who serve as a baseline for categorical variables are in the youngest age group (18 - 24), income is below 25 percentile, education below high school, being unemployed or do not know the industry that they are in, living in NSW, without voting preference, and indifferent policy satisfaction. Full results are available in Appendix Table 2. \* = p value<0.10; \*\* = p value<0.05; \*\*\* = p value<0.01.

Appendix Table A2 shows that males, those in older age groups, those with children under 18, those working in accommodation and food services, public admin and safety, and other services were less likely to receive any vaccine after September 30^th^: that is they were more likely to have been vaccinated earlier. There is a strong age gradient suggesting that older people were more likely to get vaccinated before October 1^st^ reflecting that these age groups were eligible to be vaccinated earlier than the younger age groups. Those in rental, hiring, and real estate services were more likely to get vaccinated after September 30^th^ compared to those who were out of the labour force.

### Characteristics for those who enter the competition

Of those who entered the competition, 60.6% had been vaccinated (either first or second dose) before the competition opened, compared to 35.5% of non-entrants. Table 3 examines the characetristics of those who are more likely to enter the competition. Males were less likely to do so compared to females. Relative to those aged 18-24, respondents aged 50-54 were more likely to enter, while those older than 65 were less likely to enter. Compared to those in the lowest income quartile, people in the highest income quartile were more likely to enter. Those working in manufacturing, electricity, gas, water services, accommodation and food services, transport, postal and warehousing, and financial and insurance services were less likely to enter than those who were unemployed. Respondents in LGAs with higher vaccination rates were more likely to enter. Compared to those living in NSW, respondents living in Victoria, Queensland, and Western Australia were more likely to enter M$V.

**Table 3:** Association with entry into M$V (n=2,362)

|  | Odds ratio | 95% CI |  |
| --- | --- | --- | --- |
| Male | 0.756** | 0.574 | 0.994 |
| Age 25 – 34 | 1.007 | 0.624 | 1.624 |
| Age 35 – 44 | 1.230 | 0.750 | 2.019 |
| Age 45 – 49 | 1.294 | 0.736 | 2.274 |
| Age 50 – 54 | 1.860** | 1.070 | 3.235 |
| Age 55 – 64 | 1.316 | 0.799 | 2.167 |
| Age 65 – 74 | 0.534* | 0.285 | 1.003 |
| Age 75 above | 0.145*** | 0.055 | 0.381 |
| Having a child under 18 | 0.891 | 0.652 | 1.216 |
| High school graduated | 0.816 | 0.509 | 1.309 |
| Some college | 1.078 | 0.705 | 1.648 |
| University and above | 1.280 | 0.821 | 1.994 |
| Income: 25 - 50 percentile | 1.339 | 0.871 | 2.060 |
| Income: 50 - 75 percentile | 1.317 | 0.827 | 2.097 |
| Income: 75 percentile and above | 1.531 | 0.913 | 2.568 |
| Income: refused | 1.987** | 1.123 | 3.515 |
| Industry: agriculture, forestry and fishing | 0.647 | 0.237 | 1.765 |
| Industry: mining | 0.917 | 0.184 | 4.581 |
| Industry: manufacturing | 0.529 | 0.227 | 1.233 |
| Industry: electricity, gas, water and waste services | 0.151* | 0.020 | 1.153 |
| Industry: construction and wholesale | 0.885 | 0.465 | 1.685 |
| Industry: retail trade | 1.085 | 0.674 | 1.746 |
| Industry: accommodation and food services | 0.448* | 0.187 | 1.076 |
| Industry: transport, postal and warehousing | 0.212*** | 0.066 | 0.682 |
| Industry: media and telecommunication | 0.699 | 0.319 | 1.531 |
| Industry: financial and insurance services | 0.430 | 0.185 | 1.002 |
| Industry: rental, hiring, and real estate services | 0.487 | 0.098 | 2.417 |
| Industry: professional, scientific and technical | 0.700 | 0.356 | 1.376 |
| Industry: administrative and support services | 0.742 | 0.331 | 1.667 |
| Industry: public administration and safety | 0.997 | 0.460 | 2.161 |
| Industry: education and training | 0.839 | 0.434 | 1.623 |
| Industry: health care and social assistance | 0.830 | 0.495 | 1.391 |
| Industry: arts and recreation services | 1.390 | 0.469 | 4.121 |
| Industry: other services | 0.625* | 0.357 | 1.094 |
| Living in rural | 1.095 | 0.830 | 1.445 |
| VIC | 1.703*** | 1.208 | 2.401 |
| QLD | 1.668** | 1.038 | 2.680 |
| SA | 1.363 | 0.797 | 2.330 |
| WA | 2.170*** | 1.277 | 3.685 |
| ACT, TAS, NT | 1.192 | 0.620 | 2.293 |
| Fully vaccinated rate by LGA | 1.017*** | 1.004 | 1.030 |
| With financial stress | 1.101 | 0.836 | 1.450 |
| Satisfied with policy | 0.973 | 0.715 | 1.326 |
| Not satisfied with policy | 0.744* | 0.528 | 1.049 |
| Voting liberal or national | 1.032 | 0.719 | 1.482 |
| Voting labour | 1.112 | 0.790 | 1.564 |
| Voting greens or democrats | 0.712 | 0.441 | 1.149 |
| Wave 45 (15 - 19 Nov, 2021) | 1.055 | 0.812 | 1.371 |
| Constant | 0.034*** | 0.010 | 0.116 |

Notes: Results are based on logistic regressions and the estimates are all weighted. Respondents who serve as a baseline for categorical variables are in the youngest age group (18-24), income is below 25 percentile, education below high school, being unemployed or do not know the industry that they are in, living in NSW, without voting preference, and indifferent policy satisfaction. \* = p value<0.10; \*\* = p value<0.05; \*\*\* = p value<0.01.

## Discussion

This study finds evidence of a statistically significant association between entry into the M$V competition and receipt of vaccination after the competition opened on October 1^st^. The association was driven by those who had received a second dose after September 30^th^. Those who received their second dose after the competition opened included those who had previously received the first dose sometime before October 1^st^ and decided to schedule their second dose in response to the financial incentives. Some in this group could have brought their appointment forward or were persuaded not to delay their appointment any further. However, others in this group would not have been influenced by financial incentives if their second appointment had already been booked. This could lead to an overestimate of the effect of competition entry participation on vaccination rates.

Distinguishing between the effect of financial incentives on first and second doses is important for policy as they imply different objectives and the targeting of policy towards different groups of the population. M$V aimed to encourage the population to achieve second-dose vaccination targets more quickly than would otherwise have happened. M$V was therefore focused on individuals who are already motivated. It is not surprising that the competition was not associated with an increase in first doses given the more complex range of factors influencing vaccine hesitancy.

Our research adds to the literature using a unique and representative sample of individuals from Australia when the M$V competition was open. Previous evidence from the U.S., including several evaluations of the Ohio vaccine incentives, shows mixed results using difference-in-difference study designs. Of five studies that examined first doses^[4] [7] [8] [9] [11]^, three found evidence of an effect of incentives. ^[8] [9] [11]^ Of two studies that examined second doses ^[4] [5]^, only one found an effect.^[5]^ Two studies^[6] [10]^ used the total rate of vaccinations combining first and second doses and one of these found an effect^[10]^.

The Ohio incentives and M$V were designed differently, implemented at different times during the pandemic, and may have had different marketing campaigns and this may influence the results. The whole population of Ohio was eligible to win whereas the M$V competition required individuals to enter. In the U.S. at that time the rate of vaccination was slowing, suggesting a lack of motivation in the population. In addition, October 2021 was a time when vaccination rates were steadily increasing and when vaccination targets focussing on second doses had been set by some states that were linked to the lifting of harsh lockdowns. Generally, the Australian population was more motivated to get vaccinated and the M$V competition added to this motivation. People who were already fully vaccinated may have interpreted the competition as a reward for their patience during lockdowns and for their earlier decision to get vaccinated, and for this group therefore the competition did not influence their decision to get vaccinated.

Our results found that those with higher incomes were more likely to participate in the M$V competition. Though the literature on cash lotteries suggests those on lower incomes are more likely to enter, recall that vaccination competitions are not lotteries as they do not involve gambling.^[18][19]^ The financial incentives offered through entry into M$V were likely to have been perceived as a reward for getting vaccinated and this perception may have been more widely held by those with higher incomes. The results also showed that those in LGAs with higher vaccination rates were more likely to enter the competition compared to LGAs with lower vaccination rates, suggesting that those who might have already been vaccinated before October 1^st^ were more likely to enter. The M$V marketing campaign targeted LGAs with lower vaccination rates and so assumed the campaign would be more effective in these LGAs. Our results suggest that targeted marketing to persuade people to enter a vaccine competition could be less effective in more vaccine-hesitant populations where vaccination decisions are determined by a more complex range of factors that influence access, information, and beliefs.^[20]^ In line with the objectives of M$V, vaccine competitions are more effective as ‘nudges’ for people to get their second dose more quickly.

We do not examine the overall vaccination rate but the timing of when people received their second vaccination, so our numerical results are not comparable to those from other studies that use changes over time in population vaccination rates or the number of vaccines administered. Our data are self-reported and there is a risk of over-reporting of vaccination rates due to social desirability bias. However, this is unlikely as our self-reported rate of second vaccinations of 59.9% in the sample is lower than official data at the time it was collected (77.5% on November 1^st^ and 87% on November 30^th^). This also raises concerns about the representativeness of our sample. Though our sample is representative of states and territories and uses weights based on location, gender, and age, it is from a commercial panel where respondents might be different from the general population who do not participate in commercial panel surveys in ways we do not observe that might be correlated with entry into competitions. For example, 17% of our sample participated in the M$V compared to the national estimate of 13.7%. The use of weights will ensure the sample is more representative with respect to postcode, age, gender, and state, but we recognize that the population might not be representative with respect to other variables we do not observe in the data or which are not measured for the population.

Our results are also driven by the inclusion of the unvaccinated in the denominator of the control group (non-entrants). By design, there are no unvaccinated respondents amongst lottery entrants. It is appropriate to include the unvaccinated as we report population estimates of vaccination. If we exclude the unvaccinated then this increases the probability of receiving any vaccination amongst non-entrants from 23.6 percent to 40 percent (unweighted data from Appendix Table A1) and so the difference in the percentage vaccinated compared to competition entrants falls to be close to zero. However, the inclusion of unvaccinated respondents is necessary to reflect a population estimate of the association since the unvaccinated were eligible to be vaccinated and chose not to do so, even after the competition opened.

The role of financial incentives to increase vaccination rates remains unclear.^[3] [18] [20] [21]^ Their use as nudges to speed up vaccination is likely to be effective. Policies to increase vaccination rates depend on the context and the stage of the pandemic and may interact with other strategies to increase vaccination rates, particularly in vaccine-hesitant populations where other factors are likely to matter more than financial incentives.

## Data Availability

Statistical code for the analysis s available from the Dryad repository. TTPN Survey is a proprietary data set and researchers interested in replication need to seek access to the TTPN survey by contacting the Melbourne Institute.

https://doi.org/10.5061/dryad.rv15dv495

## Funding and acknowledgments

This research was funded by the Summer Foundation (grant number: N/A) and used data from The Taking the Pulse of the Nation (TTPN) Survey run by the Melbourne Institute:

Applied Economic and Social Research, University of Melbourne. We thank Di Winkler from the Summer Foundation for comments on an earlier draft.

## Authors contributions

AS conceived of the study, secured funding, designed the survey questions, contributed to the analysis, wrote and revised the manuscript, and interpreted the results. DJ prepared the data and conducted all statistical analyses, contributed to writing and revising the manuscript, and interpreted results.

## Conflict of Interests

None declared.

## Data sharing statement

Statistical code for the analysis s available from the Dryad repository, DOI: https://doi.org/10.5061/dryad.rv15dv495. TTPN Survey is a proprietary data set and researchers interested in replication need to seek access to the TTPN survey by contacting the Melbourne Institute.

## Ethics statement

This study was approved by the University of Melbourne Faculty of Business and Economics & Melbourne Business School Human Ethics Advisory Group (Ref: 2056754.1).

## Notes

### Competing Interest Statement

The authors have declared no competing interest.

### Summary of Updates

This revision mainly explains details of those who were not participated in the competition.

## References

[1] Jarrett C, Wilson R, O’Leary M, et al. Strategies for addressing vaccine hesitancy – A systematic review. Vaccine 2015;33(34):4180–90. doi: 10.1016/j.vaccine.2015.04.040

[2] Kim HB. Financial Incentives for COVID-19 Vaccination. Epidemiology and Health 2021:e2021088. doi: 10.4178/epih.e2021088

[3] Volpp KG, Cannuscio CC. Incentives for Immunity — Strategies for Increasing Covid-19 Vaccine Uptake. New England Journal of Medicine 2021;385(1):e1. doi: 10.1056/nejmp2107719

[4] Dave D, Friedson AI, Hansen B, et al. Association Between Statewide COVID-19 Lottery Announcements and Vaccinations. JAMA Health Forum 2021;2(10) doi: 10.1001/jamahealthforum.2021.3117

[5] Lang D, Esbenshade L, Willer R. Did Ohio’s vaccine lottery increase vaccination rates? A pre-registered, synthetic control study. Journal of Experimental Political Science, 1–19. 2022. doi:10.1017/XPS.2021.32

[6] Walkey AJ, Law A, Bosch NA. Lottery-Based Incentive in Ohio and COVID-19 Vaccination Rates. JAMA 2021;326(8):766–67. doi: 10.1001/jama.2021.11048

[7] Sehgal NKR. Impact of Vax-a-Million Lottery on COVID-19 Vaccination Rates in Ohio. Am J Med 2021;134(11):1424–26. doi: 10.1016/j.amjmed.2021.06.032 [published Online First: 20210730]

[8] Barber A, West J. Conditional Cash Lotteries Increase COVID-19 Vaccination Rates. Journal of health economics 2022. Jan; 81: 102578 doi: 10.1016/j.jhealeco.2021.102578

[9] Mallow PJ, Enis A, Wackler M, et al. COVID-19 financial lottery effect on vaccine hesitant areas: Results from Ohio’s Vax-a-million program. Am J Emerg Med 2021 doi: 10.1016/j.ajem.2021.08.053 [published Online First: 20210826]80.

[10] Acharya B, Dhakal C. Implementation of State Vaccine Incentive Lottery Programs and Uptake of COVID-19 Vaccinations in the United States. JAMA Network Open 2021;4(12):e2138238. doi: 10.1001/jamanetworkopen.2021.38238

[11] Brehm M, Brehm P, Saavedra M. The Ohio Vaccine Lottery and Starting Vaccination Rates. American Journal of Health Economics 2021 doi: 10.1086/718512

[12] Thirumurthy H, Milkman K, Volpp K, et al. Association Between Statewide Financial Incentive Programs and COVID-19 Vaccination Rates. Plus One 17(3):e0263425. 2022 doi: https://doi.org/10.1371/journal.pone.0263425

[13] Australian Technical Advisory Group on Immunisation. ATAGI statement on use of COVID-19 vaccines in an outbreak setting Canberra: Australian Government; 2021 [Available from: https://www.health.gov.au/news/atagi-statement-on-use-of-covid-19-vaccines-in-an-outbreak-setting accessed March 2022 2022.

[14] Jun, Dajung, Esperanza Vera-Toscano. Australians ready to open the ‘fortress’: But there’s still a long way to go. No. 09/21. Research Insight, 2021

[15] Wilkins, Roger, Federico Zilio. How do employees feel about COVID-19 vaccination and testing mandates? No. 17/21. Research Insight, 2021

[16] Jun, Dajung, Anthony Scott. So you don’t want the COVID-19 vaccine? Here’s what research shows will change your mind. No. 03/21. Research Insight, 2021

[17] Scott, Anthony, Ou Yang. How has the Victorian outbreak of COVID-19 changed Australians’ vaccination uptake and vaccine choice? No. 08/21. Research Insight, 2021

[18] Herring M, Bledsoe T. A Model of Lottery Participation. Demographics, Context, and Attitudes. Policy Studies Journal 1994;22(2):245–57. doi: 10.1111/j.1541-0072.1994.tb01466.x

[19] Fu HN, Monson E, Otto AR. Relationships between socioleconomic status and lottery gambling across lottery types: neighborhoodllevel evidence from a large city. Addiction 2021;116(5):1256–61. doi: 10.1111/add.15252

[20] Razai MS, Chaudhry UAR, Doerholt K, et al. Covid-19 vaccination hesitancy. BMJ 2021:n1138. doi: 10.1136/bmj.n1138

[21] Wong CA, Pilkington W, Doherty IA, et al. Guaranteed Financial Incentives for COVID-19 Vaccination: A Pilot Program in North Carolina. JAMA Intern Med 2021 doi: 10.1001/jamainternmed.2021.6170 [published Online First: 20211025]

