## supplemental Table 1 for "An observational study of the association between COVID-19 vaccination rates and entry into the Australian ‘Million Dollar Vax’ competition"

### Appendix

### Table A1. Number of respondents in each of the categories of vaccination timing

| | Entrants | Percent | Non-entrants | Percent | Total | Percent |
|---|---|---|---|---|---|---|
| 1. Received first dose before but no second dose | 6 | 1.38% | 37 | 1.92% | 43 | 1.82% |
| 2. Received first dose before and second dose after | 122 | 28.0% | 285 | 14.80% | 407 | 17.2% |
| 3. Received first and second dose before | 258 | 59.2% | 646 | 33.54% | 904 | 38.3% |
| 4. Received first dose after but not second dose | 20 | 4.59% | 82 | 4.26% | 102 | 4.32% |
| 5. Received first dose after and second dose after | 30 | 6.88% | 88 | 4.57% | 118 | 5.00% |
| 6. Unvaccinated at survey (November) | 0 | 0.0% | 788 | 40.9% | 788 | 33.4% |
| Received any dose after October (2+4+5) | 172 | 39.4% | 455 | 23.6% | 627 | 26.5% |
| Received first dose after October (4+5) | 50 | 11.5% | 170 | 8.8% | 220 | 9.3% |
| Received second dose after October (2+5) | 152 | 34.9% | 373 | 19.4% | 525 | 22.2% |
| Total | 436 | 100.0% | 1926 | 100.0% | 2362 | 100.0% |

### Table A2. Full regression results (n=2,362)

| | Any dose after September 30th | | | First dose after September 30th | | | Second dose after September 30th | | |
|---|---|---|---|---|---|---|---|---|---|
| | Odds ratio | 95% CI | | Odds ratio | 95% CI | | Odds ratio | 95% CI | |
| Competition entrant | 2.274*** | 1.727 | 2.994 | 1.341 | 0.885 | 2.033 | 2.389*** | 1.800 | 3.169 |
| Male | 0.700*** | 0.55 | 0.891 | 0.578*** | 0.399 | 0.838 | 0.794* | 0.617 | 1.021 |
| Age 25 - 34 | 1.088 | 0.75 | 1.578 | 0.916 | 0.547 | 1.535 | 1.210 | 0.826 | 1.774 |
| Age 35 - 44 | 0.668** | 0.448 | 0.995 | 0.673 | 0.388 | 1.169 | 0.792 | 0.522 | 1.201 |
| Age 45 - 49 | 0.548** | 0.33 | 0.911 | 0.383** | 0.182 | 0.804 | 0.767 | 0.447 | 1.314 |
| Age 50 - 54 | 0.309*** | 0.18 | 0.528 | 0.191*** | 0.066 | 0.554 | 0.425*** | 0.244 | 0.740 |
| Age 55 - 64 | 0.332*** | 0.212 | 0.519 | 0.270*** | 0.138 | 0.529 | 0.454*** | 0.287 | 0.716 |
| Age 65 - 74 | 0.405*** | 0.231 | 0.711 | 0.137*** | 0.054 | 0.349 | 0.542** | 0.300 | 0.977 |
| Age 75 above | 0.229*** | 0.102 | 0.512 | 0.108*** | 0.023 | 0.515 | 0.284** | 0.118 | 0.682 |
| Having a child under 18 | 0.708** | 0.54 | 0.929 | 0.886 | 0.593 | 1.325 | 0.654*** | 0.494 | 0.868 |
| HS graduated | 1.104 | 0.72 | 1.694 | 1.296 | 0.692 | 2.425 | 0.953 | 0.603 | 1.506 |
| Some college | 0.961 | 0.655 | 1.41 | 1.059 | 0.604 | 1.857 | 0.932 | 0.624 | 1.391 |
| University and above | 0.957 | 0.632 | 1.45 | 0.908 | 0.483 | 1.706 | 0.926 | 0.599 | 1.432 |
| Income: 25 - 50 percentile | 1.122 | 0.762 | 1.65 | 1.201 | 0.689 | 2.091 | 1.135 | 0.750 | 1.718 |
| Income: 50 - 75 percentile | 1.041 | 0.693 | 1.563 | 1.043 | 0.566 | 1.921 | 1.169 | 0.758 | 1.801 |
| Income: 75 percentiles and above | 1.009 | 0.633 | 1.607 | 1.289 | 0.647 | 2.569 | 1.032 | 0.631 | 1.688 |
| Income: refused | 1.141 | 0.607 | 2.146 | 1.758 | 0.771 | 4.005 | 0.903 | 0.455 | 1.792 |
| Industry: agriculture, forestry and fishing | 2.465* | 0.995 | 6.108 | 2.843 | 0.834 | 9.693 | 1.353 | 0.507 | 3.606 |
| Industry: mining | 3.445* | 0.942 | 12.594 | 6.204*** | 1.777 | 21.652 | 0.442 | 0.076 | 2.570 |
| Industry: manufacturing | 0.946 | 0.418 | 2.138 | 1.133 | 0.363 | 3.539 | 0.803 | 0.352 | 1.832 |
| Industry: electricity, gas, water and waste service | 0.618 | 0.176 | 2.175 | 1.165 | 0.246 | 5.513 | 0.559 | 0.145 | 2.152 |
| Industry: construction and wholesale | 1.035 | 0.605 | 1.771 | 0.772 | 0.321 | 1.854 | 1.052 | 0.601 | 1.842 |
| Industry: retail trade | 1.112 | 0.719 | 1.718 | 0.775 | 0.389 | 1.544 | 1.004 | 0.641 | 1.572 |
| Industry: accommodation and food services | 0.369** | 0.161 | 0.85 | 0.472 | 0.166 | 1.341 | 0.485* | 0.209 | 1.129 |
| Industry: transport, postal and warehousing | 1.681 | 0.822 | 3.436 | 2.430* | 0.923 | 6.396 | 1.237 | 0.611 | 2.504 |
| Industry: media and telecommunication | 1.589 | 0.784 | 3.22 | 0.970 | 0.365 | 2.580 | 1.748 | 0.850 | 3.593 |
| Industry: financial and insurance services | 1.089 | 0.576 | 2.059 | 0.840 | 0.261 | 2.700 | 0.956 | 0.487 | 1.879 |
| Industry: rental, hiring and real estate services | 3.026** | 1.217 | 7.523 | 6.700** | 2.037 | 22.043 | 1.006 | 0.323 | 3.133 |
| Industry: professional, scientific and technical | 1.191 | 0.666 | 2.129 | 1.256 | 0.520 | 3.031 | 1.107 | 0.613 | 1.999 |
| Industry: administrative and support services | 0.859 | 0.429 | 1.72 | 1.209 | 0.483 | 3.029 | 0.948 | 0.470 | 1.912 |
| Industry: public administration and safety | 0.439* | 0.189 | 1.021 | 0.322 | 0.080 | 1.301 | 0.526 | 0.227 | 1.218 |
| Industry: education and training | 1.441 | 0.833 | 2.492 | 0.558 | 0.225 | 1.384 | 1.592 | 0.909 | 2.789 |
| Industry: health care and social assistance | 0.799 | 0.473 | 1.349 | 0.794 | 0.364 | 1.733 | 0.890 | 0.506 | 1.566 |
| Industry: arts and recreation services | 1.192 | 0.393 | 3.611 | 1.125 | 0.261 | 4.850 | 0.928 | 0.312 | 2.763 |
| Industry: other services | 0.547** | 0.316 | 0.947 | 0.814 | 0.350 | 1.892 | 0.564* | 0.318 | 1.001 |
| Living in rural | 1.251* | 0.963 | 1.625 | 1.296 | 0.892 | 1.882 | 1.207 | 0.915 | 1.592 |
| VIC | 1.329* | 0.99 | 1.783 | 2.458*** | 1.504 | 4.016 | 1.076 | 0.799 | 1.449 |
| QLD | 0.926 | 0.608 | 1.409 | 2.796*** | 1.535 | 5.093 | 0.605** | 0.389 | 0.942 |
| SA | 0.905 | 0.557 | 1.472 | 3.307*** | 1.705 | 6.416 | 0.616* | 0.368 | 1.033 |
| WA | 1.316 | 0.814 | 2.126 | 3.117*** | 1.601 | 6.069 | 0.847 | 0.510 | 1.405 |
| ACT, TAS, NT | 0.676 | 0.364 | 1.256 | 2.441* | 0.961 | 6.202 | 0.543* | 0.281 | 1.051 |
| Fully vaccinated rate by LGA | 1 | 0.988 | 1.013 | 1.010 | 0.995 | 1.025 | 0.998 | 0.985 | 1.010 |
| With financial stress | 0.873 | 0.675 | 1.128 | 0.832 | 0.563 | 1.229 | 0.925 | 0.708 | 1.209 |
| Satisfied with policy | 0.857 | 0.644 | 1.141 | 1.097 | 0.703 | 1.711 | 0.724** | 0.538 | 0.972 |
| Not satisfied with policy | 1.004 | 0.738 | 1.364 | 0.896 | 0.572 | 1.402 | 0.979 | 0.710 | 1.350 |
| Voting liberal or national | 1.082 | 0.764 | 1.533 | 0.998 | 0.574 | 1.735 | 1.185 | 0.822 | 1.709 |
| Voting labour | 1.366* | 0.987 | 1.89 | 1.226 | 0.754 | 1.996 | 1.386* | 0.985 | 1.949 |
| Voting greens or democrats | 1.148 | 0.772 | 1.705 | 1.101 | 0.620 | 1.955 | 1.432* | 0.952 | 2.155 |
| wave 45 (15 - 19 Nov, 2021) | 1.283** | 1.004 | 1.639 | 1.884*** | 1.322 | 2.686 | 1.216 | 0.941 | 1.572 |
| Constant | 0.433 | 0.261 | 1.411 | 0.027*** | 0.006 | 0.123 | 0.449 | 0.134 | 1.509 |

Notes: Results are based on logit regressions and are all weighted. Respondents who serve as a baseline are as follows: in the youngest age group (18 - 24), income below 25 percentile, education below high school, being out of labour force or do not know the industry that they are in, living in NSW, without voting preference, and indifferent policy satisfaction. * = p value<0.10; ** = p value<0.05; *** = p value<0.01.
